# Short-term forecasts to inform the response to the Covid-19 epidemic in the UK

**DOI:** 10.1101/2020.11.11.20220962

**Authors:** S Funk, S Abbott, BD Atkins, M Baguelin, JK Baillie, P Birrell, J Blake, NI Bosse, J Burton, J Carruthers, NG Davies, D De Angelis, L Dyson, WJ Edmunds, RM Eggo, NM Ferguson, K Gaythorpe, E Gorsich, G Guyver-Fletcher, J Hellewell, EM Hill, A Holmes, TA House, C Jewell, M Jit, T Jombart, I Joshi, MJ Keeling, E Kendall, ES Knock, AJ Kucharski, KA Lythgoe, SR Meakin, JD Munday, PJM Openshaw, CE Overton, F Pagani, J Pearson, PN Perez-Guzman, L Pellis, F Scarabel, MG Semple, K Sherratt, M Tang, MJ Tildesley, E Van Leeuwen, LK Whittles, CMMID COVID-19 Working Group, Imperial College COVID-19 Response Team, ISARIC4C Investigators

## Abstract

**Background:** Short-term forecasts of infectious disease can aid situational awareness and planning for outbreak response. Here, we report on multi-model forecasts of Covid-19 in the UK that were generated at regular intervals starting at the end of March 2020, in order to monitor expected healthcare utilisation and population impacts in real time.

**Methods:** We evaluated the performance of individual model forecasts generated between 24 March and 14 July 2020, using a variety of metrics including the weighted interval score as well as metrics that assess the calibration, sharpness, bias and absolute error of forecasts separately. We further combined the predictions from individual models into ensemble forecasts using a simple mean as well as a quantile regression average that aimed to maximise performance. We compared model performance to a null model of no change.

**Results:** In most cases, individual models performed better than the null model, and ensembles models were well calibrated and performed comparatively to the best individual models. The quantile regression average did not noticeably outperform the mean ensemble.

**Conclusions:** Ensembles of multi-model forecasts can inform the policy response to the Covid-19 pandemic by assessing future resource needs and expected population impact of morbidity and mortality.

## Introduction

Since the first confirmation of a case on 31 January 2020, the Covid-19 epidemic in the UK has caused a large burden of morbidity and mortality. Following a rapid increase in cases throughout February and March, triggered by repeated introduction and subsequent local transmission^1^, the UK population was advised on 16 March to avoid non-essential travel and contact with others, and to work from home if possible. This advice became enforceable law a week later, which was followed by a decline in reported cases and deaths starting in the first half of April. During the same period, hospitals prepared for a rapid increase in seriously ill patients by maximising inpatient and critical care capacity^2^.

Short-term forecasts of infectious diseases are increasingly being used to inform public health policy for a variety of diseases^3^. Models for short-term forecasts can be statistical (investigating the changing distribution of variables over time), mechanistic (explicitly incorporating plausible biological and social mechanisms of transmission), or a hybrid of the two. While the best choice of models for short-term infectious disease forecasts is an ongoing topic of research, there is some evidence that introducing mechanistic assumptions does not necessarily improve short-term predictive performance compared to statistical models that have no specific assumptions related to the disease transmission process^4^. While short-term forecasts are most prominent for seasonal influenza, more recently they have also been made for outbreaks such as Ebola, measles, Zika, and diphtheria ^5–9^. Developing accurate and reliable short-term forecasts in real time for novel infectious agents such as SARS-CoV-2 in early 2020 is particularly challenging because of uncertainty about modes of transmission, severity profiles and other relevant parameters^7,10^.

Here, we report on short-term forecasts of the Covid-19 epidemic produced by six groups in the UK, representing a mixture of academic and government institutions and using a variety of models and methods. A system for collating and aggregating these forecasts was set up through a short-term forecasting subgroup of the Scientific Pandemic Influenza Group on Modelling (SPI-M) on 23 March, 2020. The aim of these efforts was to predict the burden on the healthcare system, as well as key indicators of the current status of the epidemic faced by the UK at the time. Aggregate forecasts were made available to the Strategic Advisory Group of Experts (SAGE) and the UK government, marking the first time that a multitude of models for short-term forecasting models were explicitly combined to inform health policy in the UK. We review forecasts between the end of March and July and assess the quality of the predictions made at different times.

## Methods

### Targets and validation datasets

Initially seven forecasting targets were set: 1) The number of intensive care unit (ICU) beds occupied by confirmed Covid-19 patients, 2) the total number of beds (including ICU) occupied by confirmed Covid-19 patients, 3) the total number of deaths by date of death, 4) the number of deaths in hospitals by date of report, 5) the number of new and newly admitted confirmed Covid-19 patients in hospital, 6) the number of new admissions to ICU and 7) the cumulative number of infections.

Validation data sets for England and the seven English National Health Service (NHS) regions into which it is divided were derived from NHS England situational reports, and for the devolved nations in Northern Ireland, Scotland and Wales from their distinct reports and data sets. Because of differences in the structure of these reports and the data included in them, validation data sets started being used for the short-term forecasts at different times. Over time, this was reduced to four targets: the number of ICU beds and any beds occupied by confirmed Covid-19 patients, respectively; the number of new and newly admitted Covid-19 patients in hospital; and the number of deaths by date of death. Data sources and their interpretation changed at several points and definitions were updated as time progressed. Data sources that were not available at the time forecasts were made were excluded from the evaluation.

### Forecasts

Starting on 24 March 2020, modelling teams produced probabilistic forecasts for every day three weeks ahead for all targets except the cumulative number of infections, for which only a single time point at the date of report (i.e., a nowcast) was produced. Initially, teams submitted forecasts three times a week (with deadlines on Sunday, Tuesday and Thursday nights) and provided a best predicted estimate with lower and upper bounds of each forecast metric. On 31 March, this was changed to the median, 1%, 5%, 25%, 75%, 95% and 99% predictive quantiles. On 14 April it was decided to change to a schedule of submission twice a week (Sundays and Wednesdays, with an initial submission on a Thursday), and on 18 April the prediction quantiles were changed to 5% intervals from 5% to 95%. On 25 May the submission was changed to weekly on Tuesday mornings.

### Models

The number of models providing forecasts changed over time, with 11 models from six institutions used from the end of March. Some of these 11 models were only used on a few occasions owing to the time required in producing forecasts at regular time intervals along with shifting priorities. Other models were changed over time as discussions over the nature of the validation data sets evolved and more information became available about the intricacies of the data used to model the epidemic. One of the models (NHSBHM) was purely statistical, one a statistical/mechanistic hybrid (EpiSoon) and all others mechanistic. In brief, the models used were:

#### NHSBHM

A statistical Bayesian hierarchical model fitted individually to ICU and hospital admissions data to produce forecasts at the level of individual health trusts. The growth (decay) of the recorded values in each trust were assumed to follow a negative binomial distribution, whose mean was parameterised as a generalised logistic growth (decay) function. These forecasts were then aggregated to the regional and national level.

#### Microsimulation^11^

A spatial microsimulation model of Covid-19 transmission, fitted to hospital prevalence and incidence and death data by region of Great Britain via a sweep over a multidimensional parameter grid of the basic reproduction number *R*_*0*_, seeding and infection timing and effectiveness. A posterior distribution is calculated via Monte-Carlo sampling based on the (assumed negative binomial) likelihood of each model run.

#### SIRCOVID^12^

An age-structured stochastic compartmental Susceptible-Exposed-Infectious-Recovered (SEIR)-type model incorporating hospital care-pathways and transmission within care homes. A Bayesian evidence synthesis approach is applied to fit the model to multiple regional data sources, namely: daily deaths in hospital- and non-hospital settings, ICU and general bed prevalence data, Pillar 2 Polymerase Chain Reaction (PCR) testing data and serological survey data from blood-donors. Particle Markov-chain Monte Carlo (MCMC) methods are used to sample from the joint posterior distribution of model parameters, including disease-severity and time-varying transmission rates, and epidemic trajectories. The future epidemic trajectories are then simulated via posterior predictive forecasting.

#### Exponential growth/decline^13^

An exponential model based on assumed doubling/halving times, applied to daily hospital admission data to forecast future admissions, broken down by general hospitalisation and ICU. For each admission, the duration of hospitalisation is simulated from discretized Gamma distributions parameterised with published estimates of length of stay in ICU/non-ICU. Beds are counted for each day to derive bed occupancy in each.

#### EpiSoon^14^

A semi-mechanistic model that combines a time series forecasting model with an estimated trajectory of the time-varying reproduction number over time. Cases, hospitalisations and deaths are simulated forward separately using a renewal equation model. Changes in the reproduction number are estimated from probabilistic reconstruction of infection dates, separately for each geography.

#### Transmission^15^

An age-structured dynamic transmission model that uses Google mobility data to parameterize the impact of social distancing measures in each NHS region. The model is fitted to deaths and hospital bed occupancy in each region.

#### DetSEIRwithNB^16^

A deterministic SEIR-type model without age structure, but with differential rates between compartments depending on the next state individuals are progressing to (e.g. symptomatic cases recovering naturally or being admitted to hospital spend different times in their infectious state, reflecting different underlying progression processes). Infected cases can be asymptomatic or symptomatic, symptomatic cases recover or go to hospital, hospitalised cases recover, die or proceed to ICU, and ICU cases die or step down to hospital and then recover. The model fits new and newly confirmed cases in hospital, hospital and ICU beds occupied, and hospital deaths, using a negative binomial likelihood around the deterministic mean, to data for each region and nation independently. Transmission, and hence the reproduction number, are piecewise constant, with change points at the time of the lockdown (24 March), 1 month later (suggested by visual inspection of the data stream) and 6 weeks before the most recent data point. Two different variants for the fitting procedures are employed, one based on Maximum Likelihood Estimation (MLE) and one on MCMC.

#### Regional/age^17^

An adaptation of a Bayesian modelling framework developed for pandemic influenza^18^, this approach combines parallel deterministic SEIR transmission and disease reporting models for each NHS region, fitted to age-specific death and serological data. The parallel regions are linked through common parameters for the infectious period and the IFR. The model outputs Bayesian posterior probability distributions for parameters of interest and predictive distributions for the trajectory of the epidemic.

#### Secondary care ABC

A model which accounts for individuals admitted to ICUs and general hospital admissions, taking into consideration the potentially different timescales of fatality and recovery, fitted to hospital deaths, hospital admissions, ICU prevalence and hospital prevalence using Approximate Bayesian Computation Sequential Monte Carlo (ABC SMC).

#### StructuredODE^19,20^

An age-structured model based on SEIR-type equations but extended to include symptomatic and asymptomatic individuals, and to account for household isolation and quarantining. Susceptibility, risk of symptoms, risk of hospitalisation and risk of death are all age-dependent and based on reported data for England. For the seven regions in England and the three devolved nations (Wales, Scotland and Northern Ireland) solutions are matched to number of deaths (using date of death), hospital occupancy, ICU occupancy and hospital admissions; for England this is supplemented with serological data from blood donor sampling - this fitting procedure infers early growth and the strength of controls in each area as well as the appropriate scaling between symptomatic cases and observable health-care metrics.

### Assessment metrics

We assessed weekly forecasts using the weighted average interval score (WIS) across all quantiles that were being gathered^21^. This WIS is a strictly proper scoring rule, that is, it is optimised for predictions that come from the data-generating model and, as a consequence, encourages forecasters to report predictions representing their true belief about the future^22^. The WIS represents a parsimonious approach to scoring forecasts when only quantiles are available:

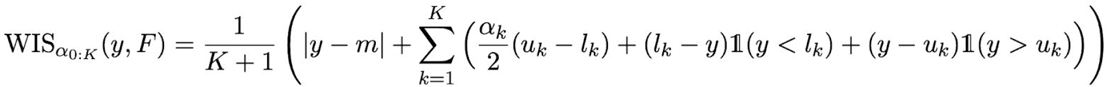

It is a weighted average of the interval score^22^ over *K* central (1 − *α*)prediction intervals bounded by quantile levels{*α*_*k*_/2, 1 − *α*_*K*_ /2}, where *y* is an observed outcome, *F* the forecast, *m* the median of the predictive distribution, and *u*_*k*_ and *l*_*k*_ are the predictive upper and lower quantiles corresponding to the central predictive interval level *k*, respectively. The interval score is optimised at 0, representing a point forecast which is exactly correct. It penalises wide prediction intervals as well as data that lies outside the intervals.

We further assessed the calibration, sharpness and bias of forecasts separately, in line with the premise that forecasts should “maximise sharpness subject to calibration”, that is as narrow as possible while consistent with future observations^22,23^. For calibration, we assessed coverage at the central 50% and 90% prediction intervals. For sharpness metric, we used the sharpness term in the WIS definition,

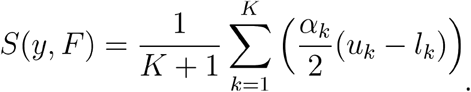

As bias metric, we estimates the proportion of predictive probability mass below/above the observation using the discrete quantiles, approximating a previously defined bias metric^23^,

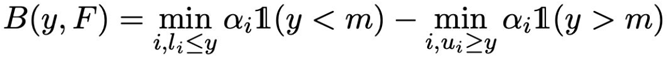

where we define the outermost quantiles corresponding to *α*_*i*_ =1 as *l*_*i*_ and *u*_*i*_ = ∞.

We lastly compared weekly forecasts by the mean absolute error (MAE) of the median forecast.

### Ensembles

We combined all the model forecasts into a single ensemble using two stacking procedures, with the aim to compare them in order to select an optimum procedure. Firstly, we produced an ensemble with equally-weighted quantiles (EWQ) by calculating each combined quantile as the mean or median of all the individual model predictive quantiles^24,25^. Secondly, we generated an ensemble using quantile regression averaging (QRA) to calculate each combined quantile as a weighted average of the individual model quantiles for each location and metric, where weights were estimated from past data to optimise past performance with respect to the one-week ahead WIS^26,27^. We tested a range of past data from 1 to 5 weeks to include in QRA, and several variants of this procedure, including constraining the weights to be non-negative and sum up to one or not, estimating weights per quantile (with an additional constraint to avoid quantile crossing) or one weight across quantiles, estimating common weights for NHSE regions or separate weights, and having an intercept in the regression or not. Weights were calculated using the *quantgen* R package^28^.

### Null model

We compared the performance of both the individual models and the ensembles with a null model that assumed that each target would stay at its current value indefinitely into the future, with uncertainty levels given by a discretised truncated normal distribution with lower bound 0 and a standard deviation given by past one-day ahead deviations from the value of the metric.

## Results

The initial forecasts were made just before the peak in confirmed Covid-19 hospital occupancy in early April. The 11 models were used to provide a total of 71,887 predicted days across the four final forecast targets and different geographies, during the 13 weeks up to 14 July 2020 or 8 weeks up to 9 June 2020 (for new and newly confirmed patients in hospital), respectively. Overall, the individual models broadly followed the trajectory of the epidemic in their forecasts but struggled to correctly predict the timing and height of the peak in early April (Fig. 1). Almost all individual models performed consistently better than the null model (no change) in median predictions at a 2 week horizon, but less clearly so at a 1 week horizon, when a model null model of no change sometimes outperformed individual models (Fig. 2). More details of individual model performance are given in the Supplementary Table 1.

**Figure 1:**
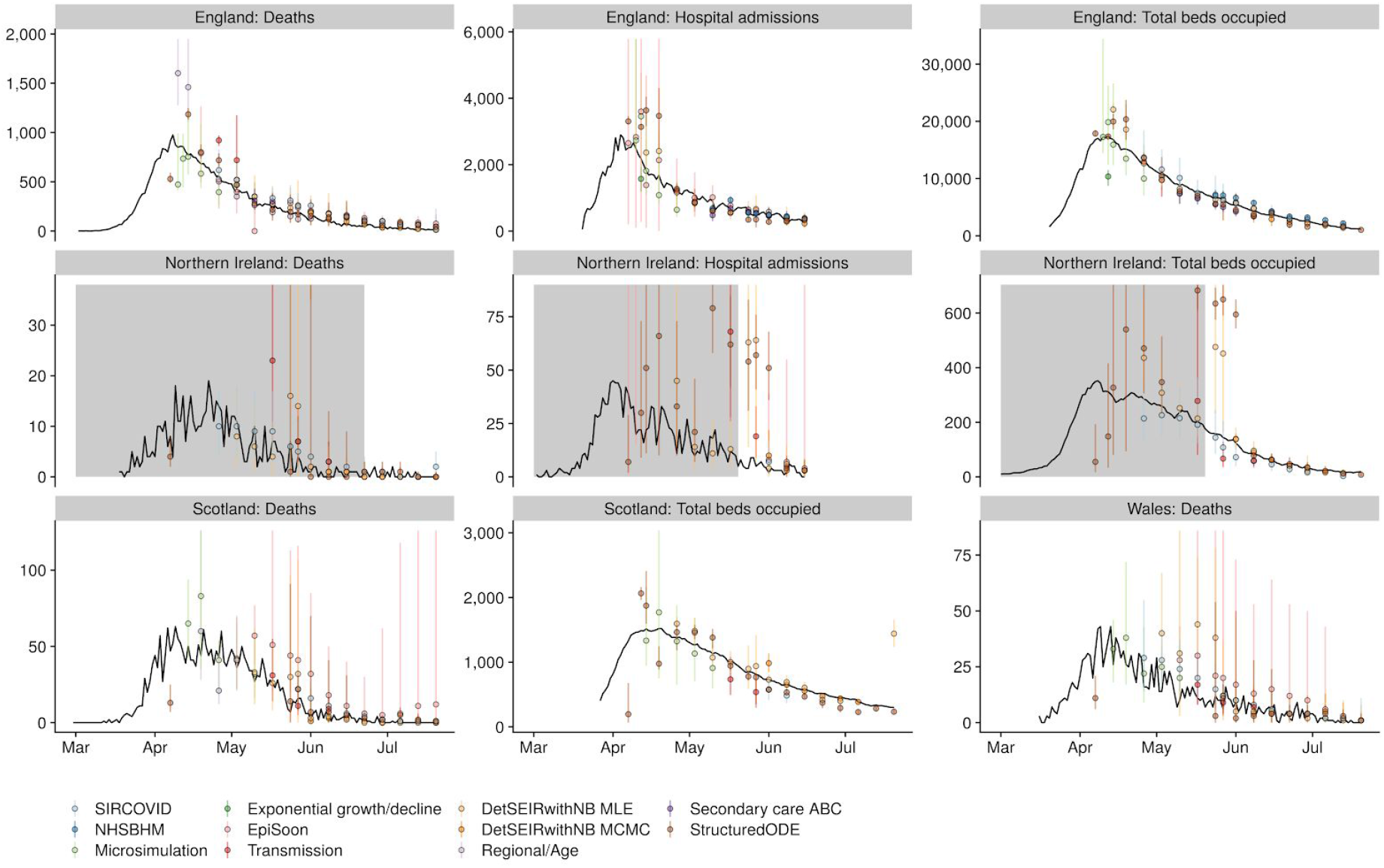
Median weekly 7-day ahead forecasts of each individual model for selected targets across the four nations of the UK (only publicly available data shown). Forecasts are shown on the day for which the forecast was made a week earlier (dots: median; whiskers: 90% prediction intervals; one colour per model) and compared to the observations (black lines). Data sources are marked in grey where they were not available at the time of the forecasts.

**Figure 2:**
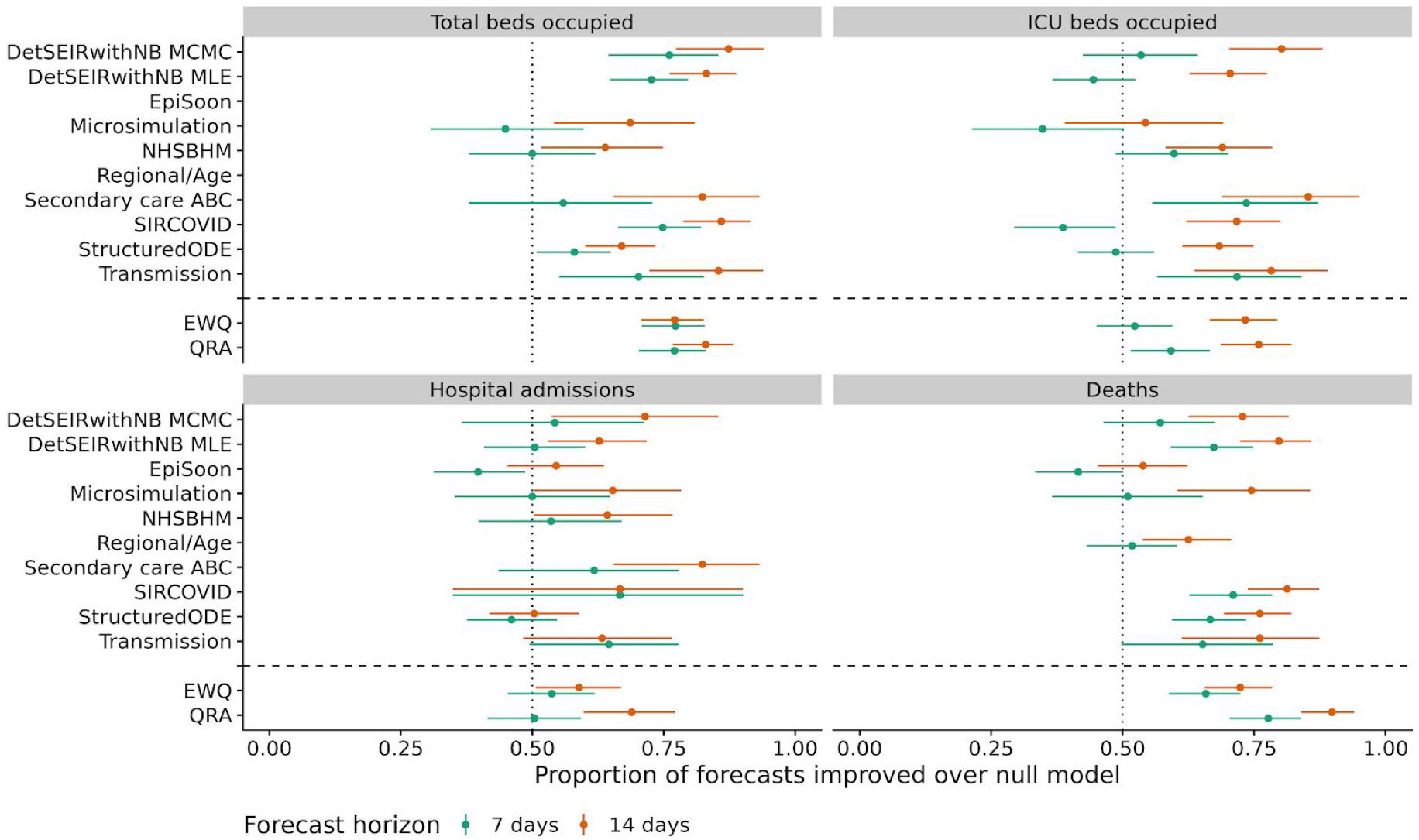
Performance against a null model of no change, shown as the proportion of 1-week and 2-week forecasts of each model that performed better than the null model with respect to the WIS (dots: proportion; lines: 95% binomial confidence intervals). Only models that were used to make forecasts at more than 2 time points are shown. Data sources are marked in grey where they were not available at the time of the forecasts.

While there was little consistency amongst the models with respect to performance against the four targets considered and no model clearly outperforming the others, all the ensemble models considered performed as good as or better than the best individual models against each target with respect to the WIS (Fig. 2 and Table 1). The best-performing QRA ensemble according to the WIS used only the latest set of historical forecasts to estimate the weights and assigned separate weights for each quantile and each geographical region, constrained to sum up to 1 and without intercept (Supplementary Table S2). The best-performing EWQ combined models by taking the median of quantiles at each quantile level (Supplementary S3). Compared to the equal-weighted ensemble, the best-performing QRA yielded some improvement (Figs. 2 and 3 and Table 1).

**Table 1:** Performance of the ensemble models (n: number of weeks for which forecasts were generated) with respect to calibration, shown as the coverage at the 50% and 90% level (Cov 0.5 and Cov 0.9, respectively), bias, sharpness (Sharp), WIS and MAE of the median. A well calibrated model would have 0.5 coverage at the 50% level and 0.9 coverage at the 90% level and a bias of zero. Models making narrower predictions have lower values of the sharpness metric, and models that are closer to the truth have lower WIS and MAE.

|  |  | 7 days ahead |  |  |  |  |  | 14 days ahead |  |  |  |  |  |
| --- | --- | --- | --- | --- | --- | --- | --- | --- | --- | --- | --- | --- | --- |
| Model | n | Cov 0.5 | Cov 0.9 | Bias | Sharp | WIS | MAE | Cov 0.5 | Cov 0.9 | Bias | Sharp | WIS | MAE |
| Total beds occupied |  |  |  |  |  |  |  |  |  |  |  |  |  |
| EWQ | 17 | 0.51 | 0.80 | -0.07 | 970 | 150 | 190 | 0.48 | 0.79 | 0.00 | 1200 | 200 | 250 |
| QRA | 17 | 0.48 | 0.82 | -0.11 | 910 | 130 | 160 | 0.43 | 0.75 | -0.08 | 1000 | 180 | 220 |
| ICU beds occupied |  |  |  |  |  |  |  |  |  |  |  |  |  |
| EWQ | 17 | 0.27 | 0.69 | 0.56 | 210 | 40 | 50 | 0.29 | 0.66 | 0.59 | 240 | 48 | 61 |
| QRA | 17 | 0.41 | 0.76 | 0.40 | 170 | 31 | 39 | 0.36 | 0.72 | 0.41 | 170 | 32 | 43 |
| Hospital admissions |  |  |  |  |  |  |  |  |  |  |  |  |  |
| EWQ | 13 | 0.42 | 0.77 | 0.41 | 520 | 86 | 110 | 0.45 | 0.77 | 0.34 | 700 | 78 | 85 |
| QRA | 13 | 0.50 | 0.86 | 0.32 | 600 | 64 | 85 | 0.42 | 0.80 | 0.19 | 540 | 54 | 68 |
| Deaths |  |  |  |  |  |  |  |  |  |  |  |  |  |
| EWQ | 18 | 0.64 | 0.93 | 0.13 | 100 | 13 | 16 | 0.66 | 0.92 | 0.18 | 130 | 15 | 18 |
| QRA | 18 | 0.55 | 0.89 | 0.02 | 72 | 11 | 13 | 0.55 | 0.89 | 0.04 | 82 | 11 | 14 |

**Figure 3:**
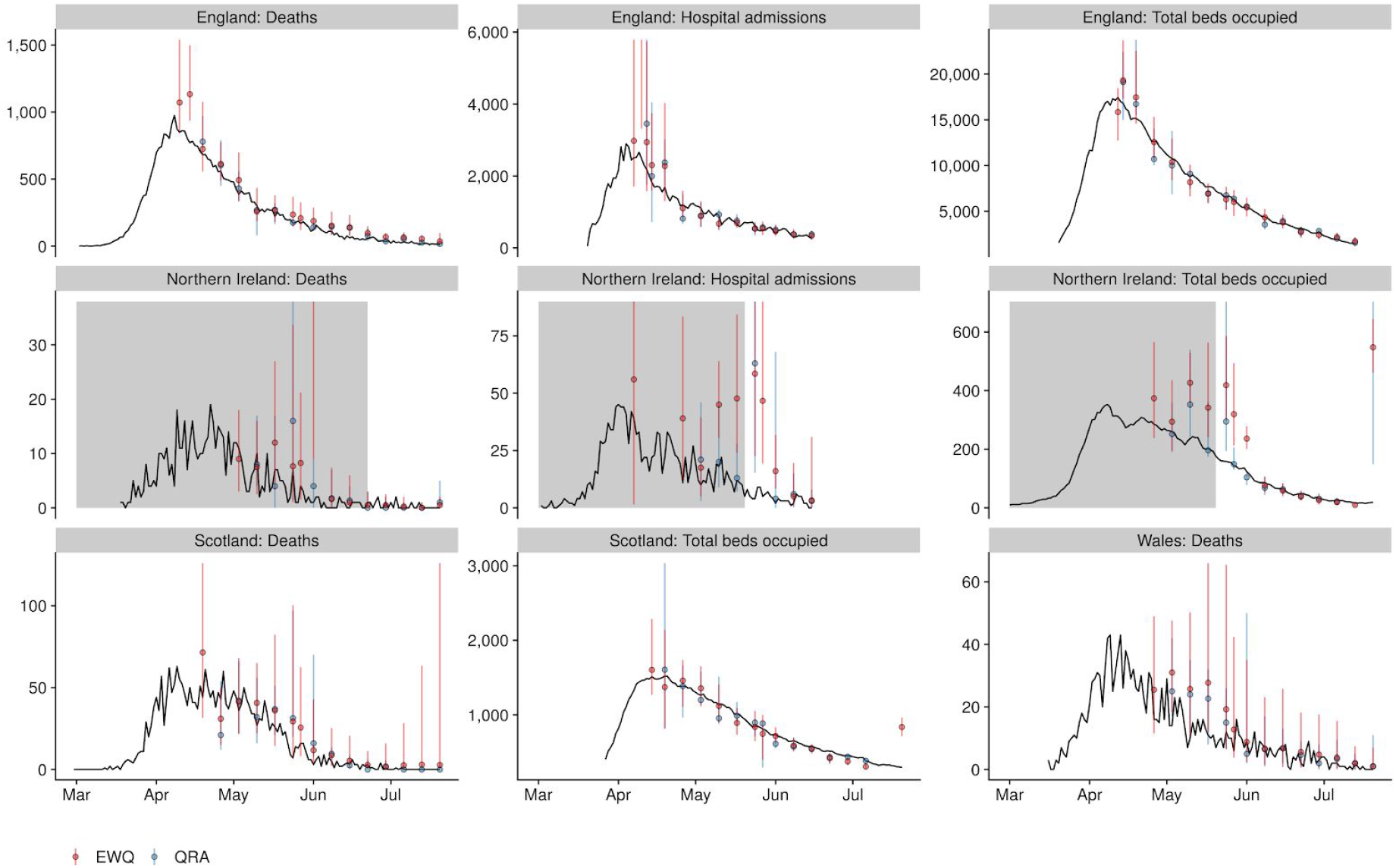
Weekly 7-day ahead forecasts of the ensemble models for selected targets across the four nations of the UK (only publicly available data shown, and only ensembles comprising more than one model). Forecasts are shown on the day for which the forecast was made a week earlier (dots: median; whiskers: 90% prediction intervals) and compared to the data as subsequently observed (black lines). Data sources are marked in grey where they were not available at the time of the forecasts.

Both the ensemble methods yielded well-calibrated models for daily deaths and new and newly confirmed cases in hospital at both 1-week and 2-week time horizons, but less so for hospital and in particular ICU occupancy (Table 1, Cov 0.5 and Cov 0.9). The forecasts were positively biased, i.e. overestimated ICU beds occupied as well as new and newly confirmed cases in hospital, while they were much less biased in either direction for total beds occupied and deaths (Table 1, Bias). The ensemble methods had similar sharpness, with the QRA model slightly sharper than the EWQ in most cases (Table 1, Sharp). Overall, the EWQ models performed better than most variants of the QRA in terms of both WIS and MAE, but the best QRA models tended to outperform the EWQ (Table 1, WIS and MAE, and Supplementary Tables 2 and 3). The QRA as a model that could learn from past performance, gave widely fluctuating weights to models over time (Fig. 4).

**Figure 4:**
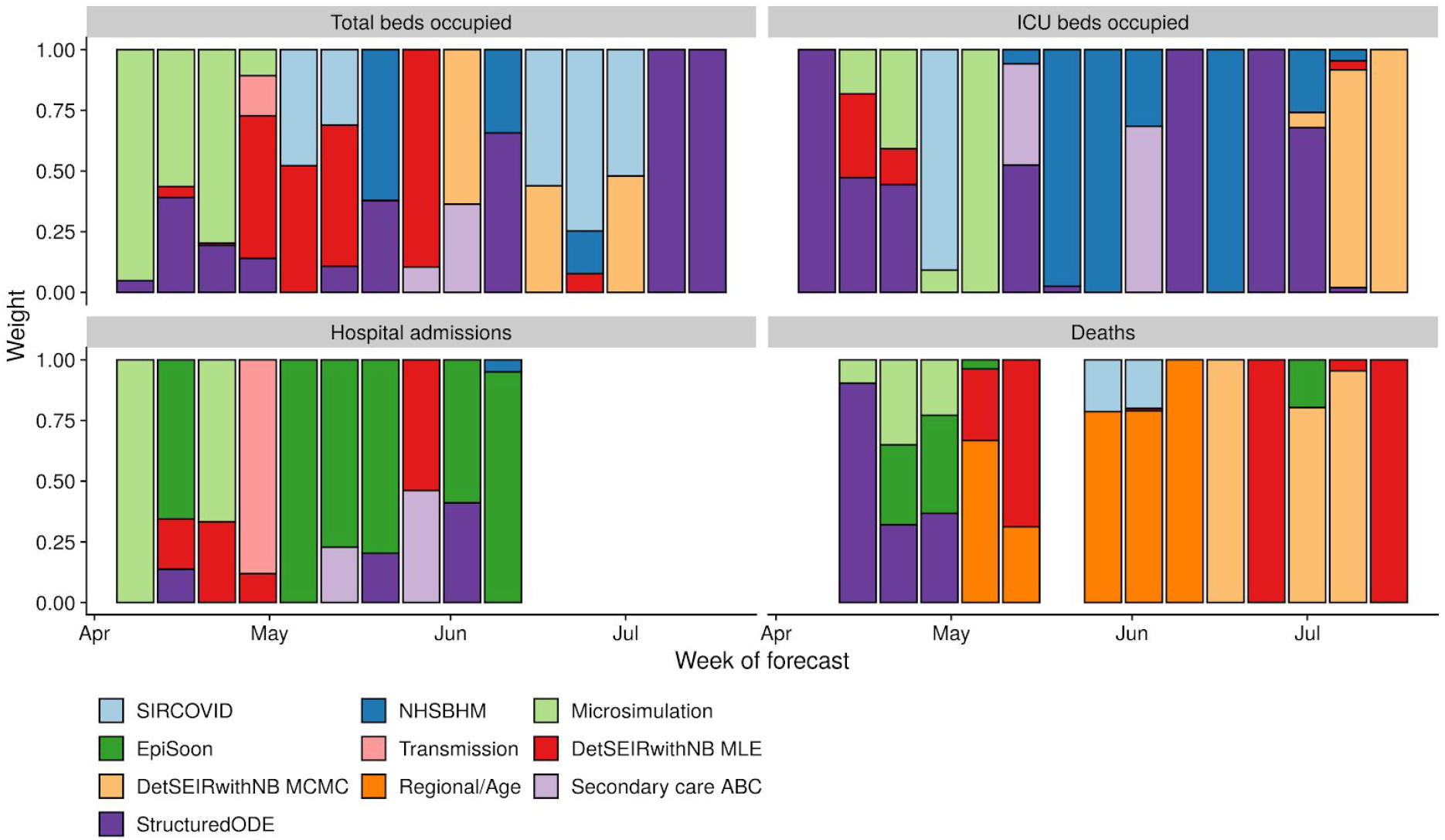
Weights given to the different models in the median prediction of the best-performing QRA at each forecast date for England.

## Discussion

A system for collating and combining short-term forecasts was set up during the early phase of the Covid-19 epidemic in the UK as a response to an urgent need for predictions of the epidemic trajectory and health system burden. This led to rapid model building and left little time for systematic testing. The results shown here highlight some of the challenges in predicting an emerging epidemic, where calibration and predictive performance are difficult to achieve and considerable uncertainties exist ^10,23,29^. The forecasts reflect a large variation in models that had been designed for a variety of purposes and, at least in some cases, rapidly adapted to the task. The models have been and are being developed and improved as additional information becomes available, both on the validation data sources and the properties of the epidemic. This makes it difficult to create like-for-like comparisons across different dates at which forecasts were made.

Given these difficulties, it is important that models are synthesised in a meaningful and principled manner in order to derive greater value from different methodological perspectives on the epidemic. We found that stacking the models using a quantile regression average that optimises historical forecast performance resulted in good calibration against most targets and tended to outperform the equally-weighted quantile combination. Having said this, this result was based on testing a wide range of ways to combine models in a quantile regression, and only the best-performing variants performed better than a simple equal-weighted quantile average, which in turn performed better than most individual models. It has been observed in other fields that a simple model average tends to outperform individual models, which can, to some degree, be explained theoretically^30^.

Over the three months analysed here, the models, as well as the inclusion and interpretation of the different data sources, were undergoing continuous change. At the same time, not every model was submitted every week due to the time pressures involved and shifting priorities in a rapidly evolving public health emergency. The lack of significant improvement from weighting by past forecast performance would indicate that these issues change the performance of the individual models on a week-by-week basis, potentially to a degree that reduces the expected benefits from systematically taking into account the past performance of each model. For these reasons, the model combination produced from the equally weighted quantiles can serve as a good and principled ensemble forecast, while the performance of different ensemble methodologies remains an ongoing topic of investigation.

Throughout the period investigated in this study, the epidemic in the UK steadily declined, and good performance in this period need not correlate with good performance in other regimes, such as a resurgence of cases or a steady-state behaviour. The difficulty of the models to correctly predict the turnaround of the epidemic around the peak (albeit often acknowledging their own uncertainty) indicates that there may be value in incorporating external information, e.g. from changing social contact studies or behavioural surveys^31,32^. All models received some weight in the QRA at least during some periods, indicating that there is value in the contributions of all of the models. Extensions to the simple regression used here^33^, or alternatives such as isotonic distributional regression^34^, may yield future performance gains, as may the inclusion of model types and structures that are currently not represented in the pool of models that are part of the ensemble.

Much discussion on real-time modelling to inform policy has focused on the value of the reproduction number, *R*. A real-time estimate of *R* indicates whether new infections are expected to increase or decrease and is, therefore, a valuable quantity to signal the need for control measures and their required strength. However, it does not provide a direct prediction of the estimated burden on the healthcare system or expected morbidity and mortality in the near future. By forecasting these directly, decision makers can be equipped with a more complete set of indicators for short-term planning than through considering *R* alone. Unlike scenario models, which are a key tool for long-term planning but are difficult to validate rigorously because of unsurmountable long-term uncertainty, modelling for short-term forecasts can be numerically evaluated and, consequently, improved in real-time. Conversely, models that are optimised for short-term forecasts usually decline in predictive performance after only a few generations of transmission^23^. Together, these two types of modelling have played a key role in informing the response to the Covid-19 pandemic in the UK and elsewhere.

As SARS-CoV-2 continues to spread in populations around the world, real time modelling and short-term forecasting can be key tools for short-term resource planning and pandemic management. The short-term forecasts described here and related initiatives in the US^35^, Germany and Poland^36^, and elsewhere are reflecting efforts to provide decision makers with the information they need to make informed decisions. In the UK, similar methodologies to the ones presented here are now used to generate medium-term projections, that is extrapolations over time periods longer than three weeks of what would be expected to happen if nothing changed from the current situation. As SARS-CoV-2 continues to affect populations around the world, short-term forecasts and longer-term projections can play a crucial part in real-time monitoring of the epidemic and its expected near-term impact in morbidity, healthcare utilisation and mortality.

## Data Availability

All publicly available data used in the manuscript is available at https://github.com/epiforecasts/covid19.forecasts.uk
Some of the data used for forecasts, and to generate the results in this manuscript, were shared with modellers in confidence and cannot be made available by the authors.

## Code and data availability

All code and data used to generate the results in this paper are available as an R package at https://github.com/epiforecasts/covid19.forecasts.uk.

## Acknowledgments

The approaches for scoring and creating ensembles have greatly benefited from regular exchange with groups conducting similar efforts elsewhere, in particular Evan Ray, Nicholas Reich (University of Massachusetts Amherst), Johannes Bracher and Tilmann Gneiting (Heidelberg Institute of Theoretical Studies). The authors would like to thank the SPI-M members and secretariat for discussion and support, especially Graham Medley, Tom Finnie and Alastair Ikin. This work has greatly benefited from parallel research undertaken by the Statistics and Dispersion Modelling team at DSTL, and was conducted within a wider effort to improve the policy response to COVID-19.

The following authors were part of the Centre for Mathematical Modelling of Infectious Disease (CMMID) COVID-19 Working Group. Each contributed in processing, cleaning and interpretation of data, interpreted findings, contributed to the manuscript, and approved the work for publication: Kevin van Zandvoort, Rosanna C Barnard, Yang Liu, Stefan Flasche, Naomi R Waterlow, James D Munday, Hamish P Gibbs, Yung-Wai Desmond Chan, C Julian Villabona-Arenas, Damien C Tully, Jack Williams, W John Edmunds, Anna M Foss, Rachel Lowe, Kiesha Prem, Kaja Abbas, Gwenan M Knight, Timothy W Russell, Christopher I Jarvis, Akira Endo, Katherine E. Atkins, Simon R Procter, Amy Gimma, Carl A B Pearson, Matthew Quaife, Megan Auzenbergs, Fiona Yueqian Sun, Petra Klepac, Alicia Rosello, Alicia Showering, David Simons, Georgia R Gore-Langton, Oliver Brady, Billy J Quilty, Emily S Nightingale, Frank G Sandmann, Graham Medley, Samuel Clifford.

The following funding sources are acknowledged as providing funding for the named authors: Wellcome Trust (210758/Z/18/Z, 206250/Z/17/Z); Health Data Research UK (grant: MR/S003975/1), the Medical Research Council (MC_PC 19065, MR/V009761/1); the National Institute for Health Research (NIHR) Health Protection Research Unite (HPRU) in Modelling and Health Economics, a partnership between PHE, Imperial College London and the London School of Hygiene & Tropical Medicine (grant code NIHR200908); the HPRU in the Health Protection Research Unit in Genomics and Enabling Data; the NIHR (16/137/109) using UK aid from the UK Government to support global health research; the European Union’s Horizon 2020 research and innovation programme - project EpiPose (No 101003688); the Bill & Melinda Gates Foundation (INV-003174). This research was partly funded by the Global Challenges Research Fund (GCRF) project ‘RECAP’ managed through RCUK and ESRC (ES/P010873/1); the Wellcome Trust and the Royal Society (202562/Z/16/Z, 107652/Z/15/Z); the Department of Mathematics at the University of Manchester for PhD funding; the Canadian Institute of Health Research (CIHR) 2019 Novel Coronavirus (COVID-19) rapid research program. The views expressed are those of the authors and not necessarily those of the United Kingdom (UK) Department of Health and Social Care, the National Health Service, the National Institute for Health Research (NIHR), or Public Health England (PHE).

The following funding sources are acknowledged as providing funding for the CMMID COVID-19 Working Group authors: Alan Turing Institute; BBSRC LIDP: BB/M009513/1; B&MGF: INV-003174, INV-001754, OPP1157270, OPP1180644, OPP1183986, NTD Modelling Consortium OPP1184344, OPP1191821; DHSC/UK Aid/NIHR: ITCRZ 03010; Elrha R2HC/UK DFID/Wellcome Trust/NIHR, DFID/Wellcome Trust: Epidemic Preparedness Coronavirus research programme 221303/Z/20/Z; ERC Starting Grant: #757699; European Commission: 101003688; Global Challenges Research Fund: ES/P010873/1; MRC: MR/N013638/1, MC_PC_19065, MR/P014658/1; Nakajima Foundation; NIHR: 16/136/46, 16/137/109, NIHR200929, PR-OD-1017-20002; Royal Society: Dorothy Hodgkin Fellowship, RP\EA\180004; Wellcome Trust: 206250/Z/17/Z, 206471/Z/17/Z, 208812/Z/17/Z, 210758/Z/18/Z; UK MRC: LID DTP MR/N013638/1.

This work uses data provided by patients and collected by the NHS as part of their care and support #DataSavesLives. We are extremely grateful to the 2,648 frontline NHS clinical and research staff and volunteer medical students, who collected this data in challenging circumstances; and the generosity of the participants and their families for their individual contributions in these difficult times. We also acknowledge the support of Jeremy J Farrar, Nahoko Shindo, Devika Dixit, Nipunie Rajapakse, Lyndsey Castle, Martha Buckley, Debbie Malden, Katherine Newell, Kwame O’Neill, Emmanuelle Denis, Claire Petersen, Scott Mullaney, Sue MacFarlane, Nicole Maziere, Julien Martinez, Oslem Dincarslan, and Annette Lake.

---

**Supplementary Table S1:** Individual model performance. Performance of individual models (n: number of weeks for which the model was used to contribute short-term forecasts) with respect to calibration, shown as the coverage at the 50% (Cov 0.5) and 90% level (Cov 0.9), bias, sharpness (Sharp), WIS and MAE. A well calibrated model would have 0.5 coverage at the 50% level and 0.9% coverage at the 90% level and a bias of zero. Models making narrower predictions have lower sharpness, and models that are closer to the truth have lower WIS and MAE. Note that performances between models are not directly comparable because they cover varying time periods.

|  |  | 7 days ahead |  |  |  |  |  | 14 days ahead |  |  |  |  |  |
| --- | --- | --- | --- | --- | --- | --- | --- | --- | --- | --- | --- | --- | --- |
| Model | n | Cov 0.5 | Cov 0.9 | Bias | Sharp | WIS | MAE | Cov 0.5 | Cov 0.9 | Bias | Sharp | WIS | MAE |
| Total beds occupied |  |  |  |  |  |  |  |  |  |  |  |  |  |
| SIRCOVID | 13 | 0.56 | 0.86 | 0.13 | 1100 | 120.0 | 150.0 | 0.50 | 0.82 | 0.17 | 1200 | 140.0 | 180.0 |
| NHSBHM | 9 | 0.14 | 0.28 | 0.73 | 360 | 170.0 | 200.0 | 0.07 | 0.24 | 0.80 | 440 | 270.0 | 310.0 |
| Microsimulation | 7 | 0.57 | 0.82 | -0.18 | 4100 | 450.0 | 540.0 | 0.37 | 0.80 | -0.28 | 5500 | 700.0 | 890.0 |
| Transmission | 5 | 0.42 | 0.80 | -0.16 | 1100 | 150.0 | 190.0 | 0.38 | 0.71 | -0.14 | 920 | 170.0 | 220.0 |
| DetSEIRwithNB MLE | 15 | 0.42 | 0.77 | -0.01 | 820 | 180.0 | 220.0 | 0.35 | 0.69 | -0.01 | 810 | 270.0 | 320.0 |
| DetSEIRwithNB MCMC | 6 | 0.35 | 0.83 | -0.22 | 600 | 99.0 | 130.0 | 0.36 | 0.82 | -0.18 | 540 | 96.0 | 120.0 |
| Secondary care ABC | 6 | 0.18 | 0.50 | -0.76 | 950 | 300.0 | 390.0 | 0.26 | 0.53 | -0.71 | 980 | 320.0 | 410.0 |
| StructuredODE | 18 | 0.21 | 0.42 | -0.23 | 500 | 240.0 | 270.0 | 0.22 | 0.41 | -0.24 | 700 | 360.0 | 400.0 |
| ICU beds occupied |  |  |  |  |  |  |  |  |  |  |  |  |  |
| SIRCOVID | 13 | 0.21 | 0.58 | 0.74 | 210 | 51.0 | 68.0 | 0.22 | 0.54 | 0.71 | 230 | 51.0 | 68.0 |
| NHSBHM | 11 | 0.22 | 0.47 | 0.54 | 94 | 26.0 | 34.0 | 0.10 | 0.32 | 0.58 | 100 | 38.0 | 46.0 |
| Microsimulation | 7 | 0.27 | 0.65 | 0.41 | 870 | 140.0 | 160.0 | 0.25 | 0.69 | 0.33 | 1200 | 150.0 | 190.0 |
| Transmission | 5 | 0.30 | 0.76 | 0.47 | 170 | 43.0 | 52.0 | 0.17 | 0.57 | 0.56 | 150 | 41.0 | 50.0 |
| DetSEIRwithNB MLE | 16 | 0.39 | 0.76 | 0.52 | 240 | 45.0 | 61.0 | 0.37 | 0.75 | 0.51 | 230 | 56.0 | 72.0 |
| DetSEIRwithNB MCMC | 8 | 0.45 | 0.77 | 0.44 | 82 | 17.0 | 22.0 | 0.41 | 0.81 | 0.41 | 74 | 14.0 | 18.0 |
| Secondary care ABC | 6 | 0.41 | 0.88 | 0.52 | 280 | 38.0 | 53.0 | 0.38 | 0.76 | 0.51 | 250 | 38.0 | 54.0 |
| StructuredODE | 18 | 0.23 | 0.45 | 0.44 | 110 | 49.0 | 57.0 | 0.21 | 0.49 | 0.49 | 140 | 61.0 | 72.0 |
| Hospital admissions |  |  |  |  |  |  |  |  |  |  |  |  |  |
| NHSBHM | 7 | 0.30 | 0.73 | 0.33 | 110 | 24.0 | 30.0 | 0.29 | 0.64 | 0.34 | 110 | 31.0 | 38.0 |
| Microsimulation | 7 | 0.45 | 0.80 | 0.12 | 660 | 91.0 | 110.0 | 0.37 | 0.80 | 0.03 | 840 | 120.0 | 150.0 |
| EpiSoon | 14 | 0.54 | 0.84 | 0.43 | 990 | 77.0 | 85.0 | 0.61 | 0.93 | 0.31 | 1300 | 87.0 | 85.0 |
| Transmission | 5 | 0.30 | 0.61 | 0.50 | 110 | 23.0 | 28.0 | 0.24 | 0.61 | 0.27 | 100 | 21.0 | 26.0 |
| DetSEIRwithNB MLE | 11 | 0.47 | 0.79 | 0.21 | 240 | 47.0 | 58.0 | 0.42 | 0.79 | 0.15 | 220 | 46.0 | 55.0 |
| DetSEIRwithNB MCMC | 3 | 0.48 | 0.94 | -0.24 | 100 | 15.0 | 20.0 | 0.27 | 0.94 | -0.25 | 88 | 15.0 | 22.0 |
| Secondary care ABC | 6 | 0.26 | 0.68 | 0.05 | 110 | 21.0 | 28.0 | 0.21 | 0.76 | -0.08 | 110 | 24.0 | 33.0 |
| StructuredODE | 13 | 0.23 | 0.42 | 0.24 | 140 | 91.0 | 100.0 | 0.21 | 0.50 | 0.16 | 180 | 120.0 | 130.0 |
| Deaths |  |  |  |  |  |  |  |  |  |  |  |  |  |
| SIRCOVID | 14 | 0.50 | 0.87 | 0.20 | 48 | 6.9 | 9.3 | 0.44 | 0.88 | 0.26 | 50 | 7.3 | 10.0 |
| Microsimulation | 7 | 0.45 | 0.76 | 0.05 | 190 | 27.0 | 37.0 | 0.55 | 0.88 | 0.05 | 240 | 26.0 | 35.0 |
| EpiSoon | 15 | 0.45 | 0.93 | 0.45 | 120 | 13.0 | 17.0 | 0.51 | 0.96 | 0.44 | 190 | 15.0 | 18.0 |
| Transmission | 5 | 0.37 | 0.78 | 0.55 | 90 | 26.0 | 32.0 | 0.28 | 0.65 | 0.68 | 82 | 29.0 | 35.0 |
| DetSEIRwithNB MLE | 13 | 0.64 | 0.87 | -0.17 | 78 | 8.0 | 9.3 | 0.64 | 0.86 | -0.16 | 71 | 7.3 | 8.7 |
| DetSEIRwithNB MCMC | 8 | 0.45 | 0.74 | -0.43 | 33 | 7.5 | 9.4 | 0.48 | 0.75 | -0.44 | 29 | 6.8 | 8.6 |
| Regional/Age | 19 | 0.35 | 0.71 | -0.35 | 120 | 29.0 | 34.0 | 0.34 | 0.62 | -0.37 | 130 | 28.0 | 34.0 |
| StructuredODE | 18 | 0.45 | 0.71 | -0.03 | 59 | 18.0 | 21.0 | 0.48 | 0.72 | -0.05 | 60 | 21.0 | 24.0 |

**Supplementary Table S2:**
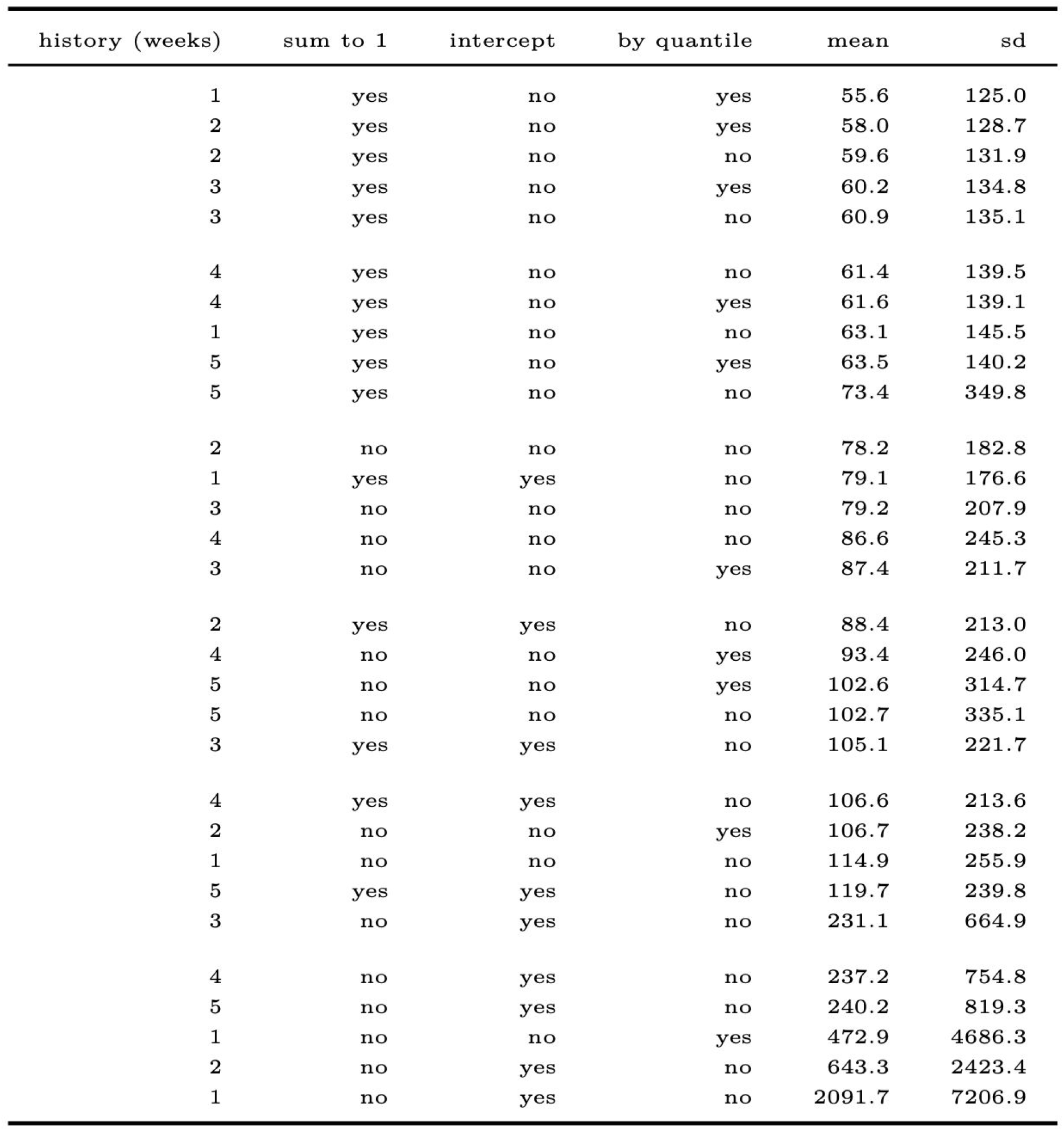
QRA ensemble performance. Performance of all permutations of QRA models with respect to the WIS. Model options were: the number of previous forecasts (history) to include in the regression, whether to force the weights to be non-negative and sum to 1, whether to model an intercept, whether to estimate separate weights by quantile, and whether to estimate separate weights by English region.

**Supplementary Table S3:** EWQ ensemble performance. Performance of mean- and median-based EWQ models with respect to the WIS.

| combination | mean | sd |
| --- | --- | --- |
| mean | 53.6 | 109.7 |
| median | 55.4 | 119.7 |

## Supplementary Text S4: ISARIC4C Investigators

*Consortium Lead Investigator:* J Kenneth Baillie.

C*hief Investigator:* Malcolm G Semple.

*Co-Lead Investigator:* Peter JM Openshaw.

*ISARIC Clinical Coordinator:* Gail Carson.

### Co-Investigators

Beatrice Alex, Benjamin Bach, Wendy S Barclay, Debby Bogaert, Meera Chand, Graham S Cooke, Annemarie B Docherty, Jake Dunning, Ana da Silva Filipe, Tom Fletcher, Christopher A Green, Ewen M Harrison, Julian A Hiscox, Antonia Ying Wai Ho, Peter W Horby, Samreen Ijaz, Saye Khoo, Paul Klenerman, Andrew Law, Wei Shen Lim, Alexander J Mentzer, Laura Merson, Alison M Meynert, Mahdad Noursadeghi, Shona C Moore, Massimo Palmarini, William A Paxton, Georgios Pollakis, Nicholas Price, Andrew Rambaut, David L Robertson, Clark D Russell, Vanessa Sancho-Shimizu, Janet T Scott, Thushan de Silva, Louise Sigfrid, Tom Solomon, Shiranee Sriskandan, David Stuart, Charlotte Summers, Richard S Tedder, Emma C Thomson, AA Roger Thompson, Ryan S Thwaites, Lance CW Turtle, Maria Zambon.

### Project Managers

Hayley Hardwick, Chloe Donohue, Ruth Lyons, Fiona Griffiths, Wilna Oosthuyzen.

### Data Analysts

Lisa Norman, Riinu Pius, Tom M Drake, Cameron J Fairfield, Stephen Knight, Kenneth A Mclean, Derek Murphy, Catherine A Shaw.

### Data and Information System Managers

Jo Dalton, James Lee, Daniel Plotkin, Michelle Girvan, Egle Saviciute, Stephanie Roberts, Janet Harrison, Laura Marsh, Marie Connor, Sophie Halpin, Clare Jackson, Carrol Gamble.

### Data integration and presentation

Gary Leeming, Andrew Law, Murray Wham, Sara Clohisey, Ross Hendry, James Scott-Brown.

### Material Management

William Greenhalf, Victoria Shaw, Sarah McDonald.

### Patient engagement

Seán Keating

### Outbreak Laboratory Staff and Volunteers

Katie A. Ahmed, Jane A Armstrong, Milton Ashworth, Innocent G Asiimwe, Siddharth Bakshi, Samantha L Barlow, Laura Booth, Benjamin Brennan, Katie Bullock, Benjamin WA Catterall, Jordan J Clark, Emily A Clarke, Sarah Cole, Louise Cooper, Helen Cox, Christopher Davis, Oslem Dincarslan, Chris Dunn, Philip Dyer, Angela Elliott, Anthony Evans, Lorna Finch, Lewis WS Fisher, Terry Foster, Isabel Garcia-Dorival, Willliam Greenhalf, Philip Gunning, Catherine Hartley, Antonia Ho, Rebecca L Jensen, Christopher B Jones, Trevor R Jones, Shadia Khandaker, Katharine King, Robyn T. Kiy, Chrysa Koukorava, Annette Lake, Suzannah Lant, Diane Latawiec, L Lavelle-Langham, Daniella Lefteri, Lauren Lett, Lucia A Livoti, Maria Mancini, Sarah McDonald, Laurence McEvoy, John McLauchlan, Soeren Metelmann, Nahida S Miah, Joanna Middleton, Joyce Mitchell, Shona C Moore, Ellen G Murphy, Rebekah Penrice-Randal, Jack Pilgrim, Tessa Prince, Will Reynolds, P. Matthew Ridley, Debby Sales, Victoria E Shaw, Rebecca K Shears, Benjamin Small, Krishanthi S Subramaniam, Agnieska Szemiel, Aislynn Taggart, Jolanta Tanianis-Hughes, Jordan Thomas, Erwan Trochu, Libby van Tonder, Eve Wilcock, J. Eunice Zhang.

### Local Principal Investigators

Kayode Adeniji, Daniel Agranoff, Ken Agwuh, Dhiraj Ail, Ana Alegria, Brian Angus, Abdul Ashish, Dougal Atkinson, Shahedal Bari, Gavin Barlow, Stella Barnass, Nicholas Barrett, Christopher Bassford, David Baxter, Michael Beadsworth, Jolanta Bernatoniene, John Berridge, Nicola Best, Pieter Bothma, David Brealey, Robin Brittain-Long, Naomi Bulteel, Tom Burden, Andrew Burtenshaw, Vikki Caruth, David Chadwick, Duncan Chambler, Nigel Chee, Jenny Child, Srikanth Chukkambotla, Tom Clark, Paul Collini, Catherine Cosgrove, Jason Cupitt, Maria-Teresa Cutino-Moguel, Paul Dark, Chris Dawson, Samir Dervisevic, Phil Donnison, Sam Douthwaite, Ingrid DuRand, Ahilanadan Dushianthan, Tristan Dyer, Cariad Evans, Chi Eziefula, Chrisopher Fegan, Adam Finn, Duncan Fullerton, Sanjeev Garg, Sanjeev Garg, Atul Garg, Effrossyni Gkrania-Klotsas, Jo Godden, Arthur Goldsmith, Clive Graham, Elaine Hardy, Stuart Hartshorn, Daniel Harvey, Peter Havalda, Daniel B Hawcutt, Maria Hobrok, Luke Hodgson, Anil Hormis, Michael Jacobs, Susan Jain, Paul Jennings, Agilan Kaliappan, Vidya Kasipandian, Stephen Kegg, Michael Kelsey, Jason Kendall, Caroline Kerrison, Ian Kerslake, Oliver Koch, Gouri Koduri, George Koshy, Shondipon Laha, Steven Laird, Susan Larkin, Tamas Leiner, Patrick Lillie, James Limb, Vanessa Linnett, Jeff Little, Michael MacMahon, Emily MacNaughton, Ravish Mankregod, Huw Masson, Elijah Matovu, Katherine McCullough, Ruth McEwen, Manjula Meda, Gary Mills, Jane Minton, Mariyam Mirfenderesky, Kavya Mohandas, Quen Mok, James Moon, Elinoor Moore, Patrick Morgan, Craig Morris, Katherine Mortimore, Samuel Moses, Mbiye Mpenge, Rohinton Mulla, Michael Murphy, Megan Nagel, Thapas Nagarajan, Mark Nelson, Igor Otahal, Mark Pais, Selva Panchatsharam, Hassan Paraiso, Brij Patel, Natalie Pattison, Justin Pepperell, Mark Peters, Mandeep Phull, Stefania Pintus, Jagtur Singh Pooni, Frank Post, David Price, Rachel Prout, Nikolas Rae, Henrik Reschreiter, Tim Reynolds, Neil Richardson, Mark Roberts, Devender Roberts, Alistair Rose, Guy Rousseau, Brendan Ryan, Taranprit Saluja, Aarti Shah, Prad Shanmuga, Anil Sharma, Anna Shawcross, Jeremy Sizer, Manu Shankar-Hari, Richard Smith, Catherine Snelson, Nick Spittle, Nikki Staines, Tom Stambach, Richard Stewart, Pradeep Subudhi, Tamas Szakmany, Kate Tatham, Jo Thomas, Chris Thompson, Robert Thompson, Ascanio Tridente, Darell Tupper-Carey, Mary Twagira, Andrew Ustianowski, Nick Vallotton, Lisa Vincent-Smith, Shico Visuvanathan, Alan Vuylsteke, Sam Waddy, Rachel Wake, Andrew Walden, Ingeborg Welters, Tony Whitehouse, Paul Whittaker, Ashley Whittington, Meme Wijesinghe, Martin Williams, Lawrence Wilson, Sarah Wilson, Stephen Winchester, Martin Wiselka, Adam Wolverson, Daniel G Wooton, Andrew Workman, Bryan Yates, and Peter Young.

## ISARIC4C funding

This work is supported by grants from: the National Institute for Health Research (NIHR; award CO-CIN-01), the Medical Research Council (MRC; grant MC_PC_19059), and by the NIHR Health Protection Research Unit (HPRU) in Emerging and Zoonotic Infections at University of Liverpool in partnership with Public Health England (PHE), in collaboration with Liverpool School of Tropical Medicine and the University of Oxford (award 200907), NIHR HPRU in Respiratory Infections at Imperial College London with PHE (award 200927), Wellcome Trust and Department for International Development (DID; 215091/Z/18/Z), the Bill and Melinda Gates Foundation (OPP1209135), Liverpool Experimental Cancer Medicine Centre (grant reference C18616/A25153), NIHR Biomedical Research Centre at Imperial College London (IS-BRC-1215-20013), EU Platform for European Preparedness Against (Re-)emerging Epidemics (PREPARE; FP7 project 602525), and NIHR Clinical Research Network for providing infrastructure support for this research. PJMO is supported by a NIHR senior investigator award (201385). The views expressed are those of the authors and not necessarily those of the Department of Health and Social Care, DID, NIHR, MRC, Wellcome Trust, or PHE.

